# Rapidly fatal pneumonitis from immunotherapy and concurrent SARS-CoV-2 infection in a patient with newly diagnosed lung cancer

**DOI:** 10.1101/2020.04.29.20085738

**Authors:** Christine M. Lovly, Kelli L. Boyd, Paula I. Gonzalez-Ericsson, Cindy L. Lowe, Hunter M. Brown, Robert D. Hoffman, Brent C. Sterling, Meghan E. Kapp, Douglas B. Johnson, Prasad R. Kopparapu, Wade T. Iams, Melissa A. Warren, Michael J. Noto, Brian I. Rini, Madan Jagasia, Suman R. Das, Justin M. Balko

## Abstract

Immune checkpoint inhibitors (ICIs) are used for the treatment of numerous cancers, but risks associated with ICI-therapy during the COVID-19 pandemic are poorly understood. We report a case of acute lung injury in a lung cancer patient initially treated for ICI-pneumonitis and later found to have concurrent SARS-CoV-2 infection. Post-mortem analyses revealed diffuse alveolar damage in both the acute and organizing phases, with a predominantly CD68+ inflammatory infiltrate. Serum was positive for anti-SARS-CoV-2 IgG, suggesting that viral infection predated administration of ICI-therapy and may have contributed to a more fulminant clinical presentation. These data suggest the need for routine SARS-CoV-2 testing in cancer patients, where clinical and radiographic evaluations may be non-specific.

## Case Report

A 56-year-old male with type 2 diabetes, chronic obstructive pulmonary disease, and ongoing tobacco use presented with a one-month history of left-sided chest pain, dyspnea, cough, and sinus congestion but no fevers or chills (**Fig. 1A**). His symptoms were refractory to a course of doxycycline. Chest CT revealed a 5 cm spiculated mass in the lingula, mediastinal lymphadenopathy, and multiple liver lesions (**Fig. 1B** and data not shown). He was admitted to the hospital for symptom management and to expedite diagnostic studies. Transbronchial biopsy of a subcarinal lymph node revealed small cell lung cancer (SCLC). He was treated with standard therapy for extensive-stage SCLC - carboplatin, etoposide, and atezolizumab (an anti-PD-L1 monoclonal antibody) then discharged on the third day of therapy. He returned to the emergency department within 48 hours with worsening dyspnea and hypoxemia but remained afebrile. Chest CT showed interval development of bilateral ground glass opacities with interlobular septal thickening, most pronounced in the right upper lobe (**Fig. 1C**), concerning for drug-induced pneumonitis versus infection. He was treated for presumed ICI-pneumonitis (from atezolizumab) with steroids (1g of methylprednisolone for 2 days followed by 1 mg/kg/day prednisone). After 5 days, his respiratory status worsened. Prednisone was increased to 2 mg/kg/day and infliximab (TNFα inhibitor, 5mg/kg) was administered. Repeat CT scan showed progression of the ground glass opacities with a decrease in the left upper lobe tumor mass (**Fig. 1D**). He continued to require 10 - 15L high-flow oxygen per nasal cannula. He was started on empiric antibiotics (vancomycin and piperacillin/tazobactam) and given one dose of intravenous immunoglobulin (1g/kg). To consider other causes for his declining respiratory status, a SARS-CoV-2 PCR test from a nasal swab was ordered, and the result was positive. Ferritin was 804 ng/mL and LDH was 1218 units/L. Steroids were weaned. His respiratory status continued to worsen, ultimately requiring intubation and mechanical ventilation for hypoxemic respiratory failure. He developed shock requiring vasopressor support. In discussion with his wife, a decision was made to transition to comfort care, and he passed away peacefully. Blood samples collected on days 22 and 30 were later analyzed for SARS-CoV-2 antibodies, both of which showed low but detectable titers of IgM and IgG to the virus receptor binding domain (**Fig. 1E,F**).

**Figure 1:**
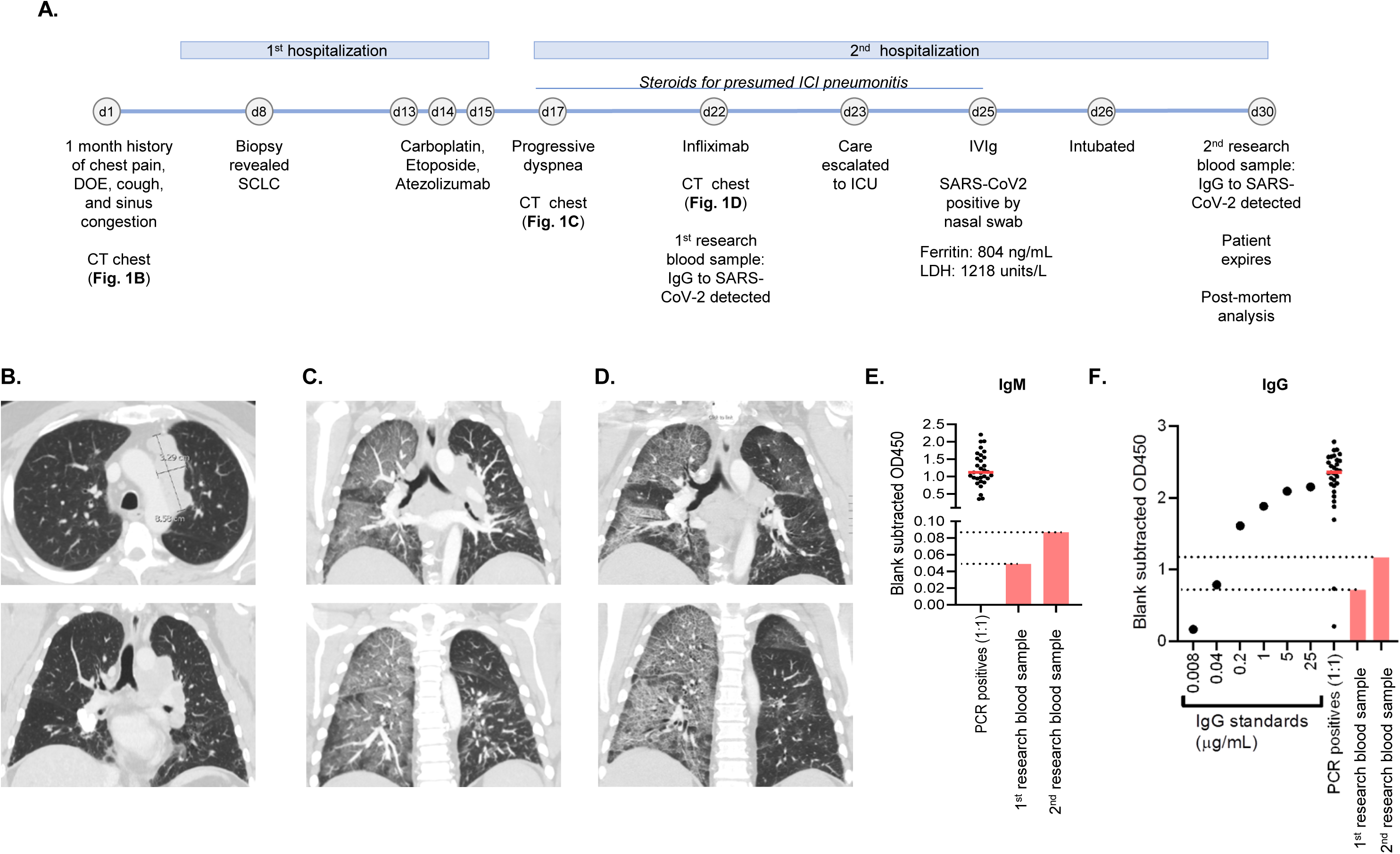
Clinical course and diagnostic studies. (**A**) Timeline depicting the index patient’s clinical course from the day of initial presentation to health care (day 0) to his demise (day 30). d: day; DOE: dyspnea on exertion; SCLC: small cell lung cancer. IVIg: intravenous immunoglobulin; LDH: lactate dehydrogenase. (**B**) CT scans of the chest on the day of the patient’s initial presentation (day 0) showing a mass in the left lung. (**C**) CT scans of the chest 2 days after completion of cycle 1 of carboplatin, etoposide, and atezolizumab demonstrating right > left ground glass opacities. (**D**) CT scans of the chest 7 days after completion of cycle 1 of carboplatin, etoposide, and atezolizumab demonstrating progression of bilateral ground glass opacities. (**E**) Direct ELISA for SARS-CoV-2 receptor binding domain specific IgM to confirm infection; 31 SARS-CoV-2 PCR positive patient serum samples were used for comparison. (**F**) Direct ELISA for SARS-CoV-2 receptor binding domain specific IgG to confirm infection; standard curve was generated using control sample provided by the kit to quantify amount of receptor binding domain specific antibody. SARS-CoV-2 PCR positive 31 serum samples were used for comparison.

## Methods

The patient consented to protocols that permitted de-identified research use of biospecimens and clinical data (IRB #030763), and family consented to a post-mortem histopathologic evaluation (IRB#191213). Tissues were fixed in 10% neutral buffered formalin and embedded in paraffin. Immunohistochemistry for CD68 (MM36–10, StatLab), CD3 (MM150–10, StatLab), CD20 (MM07–10, StatLab), Myeloperoxidase (MPO, Dako A0398), and PD-L1 (Thermo PA5-28115) was performed using the Leica Bond RX autostainer. RNAscope® with 2.5 LS Probe -V-nCoV2019-S and 2.5 LS Probe -V-nCoV2019-S-sense probes (cat.# 848568 and 845708, Advanced Cell Diagnostics) was performed on the Leica Bond RX. Multiplex immunofluorescence labeling was performed with the Ultivue UltiMapper™ I/O PD-L1 and UltiMapper™ I/O APC kits on the Leica Bond RX. Whole sections were digitally acquired using an AxioScan Z1 slide scanner (Carl Zeiss, Germany). Automated quantification was performed on whole slide images using QuPath software^1^. Plasma samples were tested for IgM and IgG antibody content at 1:1 and 1:100 dilutions using the Genscript SARS-CoV-2 Spike S1-Receptor Binding Domain IgM & IgG ELISA Detection Kit according to the manufacturers protocol (cat.# L00831). ELISA assays were performed in duplicate and repeated two independent times.

## Results

Microscopic evaluation revealed bilateral extensive diffuse alveolar damage in a predominantly organizing phase; however, the right lung had more involvement (**Fig. 2A, Fig. S1A-C**). Alveolar septa were thickened and alveolar spaces contained foamy macrophages, sloughing epithelial cells, fibromyxoid-organizing exudates, fibrin, hemorrhage, and edema (**Fig. S1A)**. There were interwoven areas mature fibrosis and fibrotic nodules in the alveolar spaces (**Fig. S1B)**. Reactive sloughed epithelial cells had amphophilic cytoplasm, enlarged nuclei, multi-nucleation and nucleoli (**Fig. S1C**).

**Figure 2:**
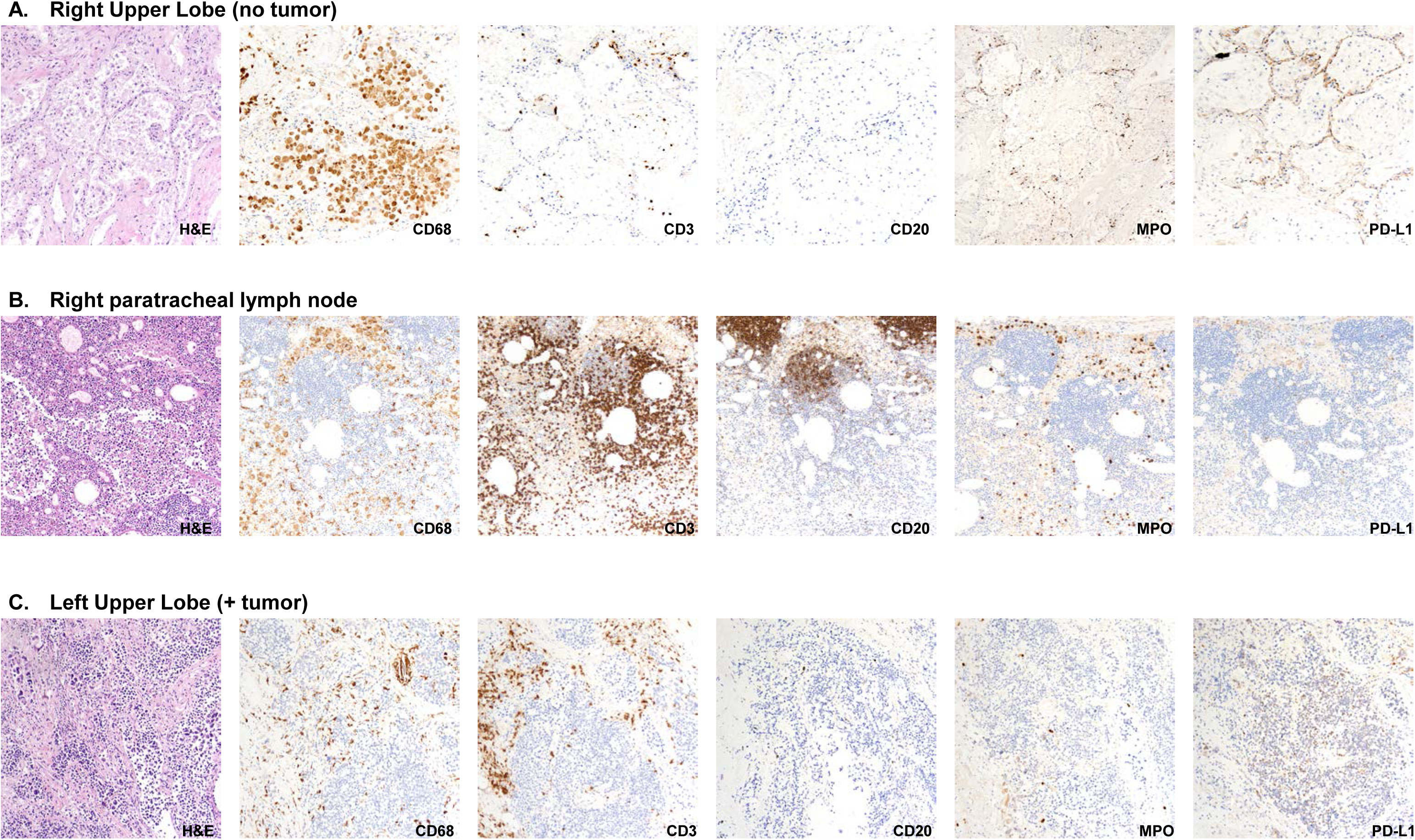
Diffuse acute lung injury with monocytic response in lung infected with SARS-CoV-2. Representative H&E images (200x magnification) showing autopsy specimens from the right upper lobe of the lung (**A**), right paratracheal lymph node (**B**), and left upper lobe of the lung (**C**). Representative immunohistochemical stains for CD68, CD3, CD20, myeloperoxidase (MPO), and PD-L1 are shown for each anatomical location.

Immunohistochemical evaluation of the inflammatory response in the pulmonary interstitium revealed that most infiltrating cells in the alveolar spaces labeled positive for CD68 (**Fig. 2A**) with a relative paucity of CD3, CD20, and myeloperoxidase (MPO) positive cells. Unlike the CD68 staining, CD3 and MPO signals were predominantly detected in the alveolar wall. CD68+ cells filled and expanded sinusoids of the draining right paratracheal lymph node (**Fig. 2B**). PD-L1 was expressed in epithelial cells, some infiltrating macrophages, and cells appearing morphologically as endothelial cells (**Figs. 2A, 3D, 3G)**.

**Figure 3:**
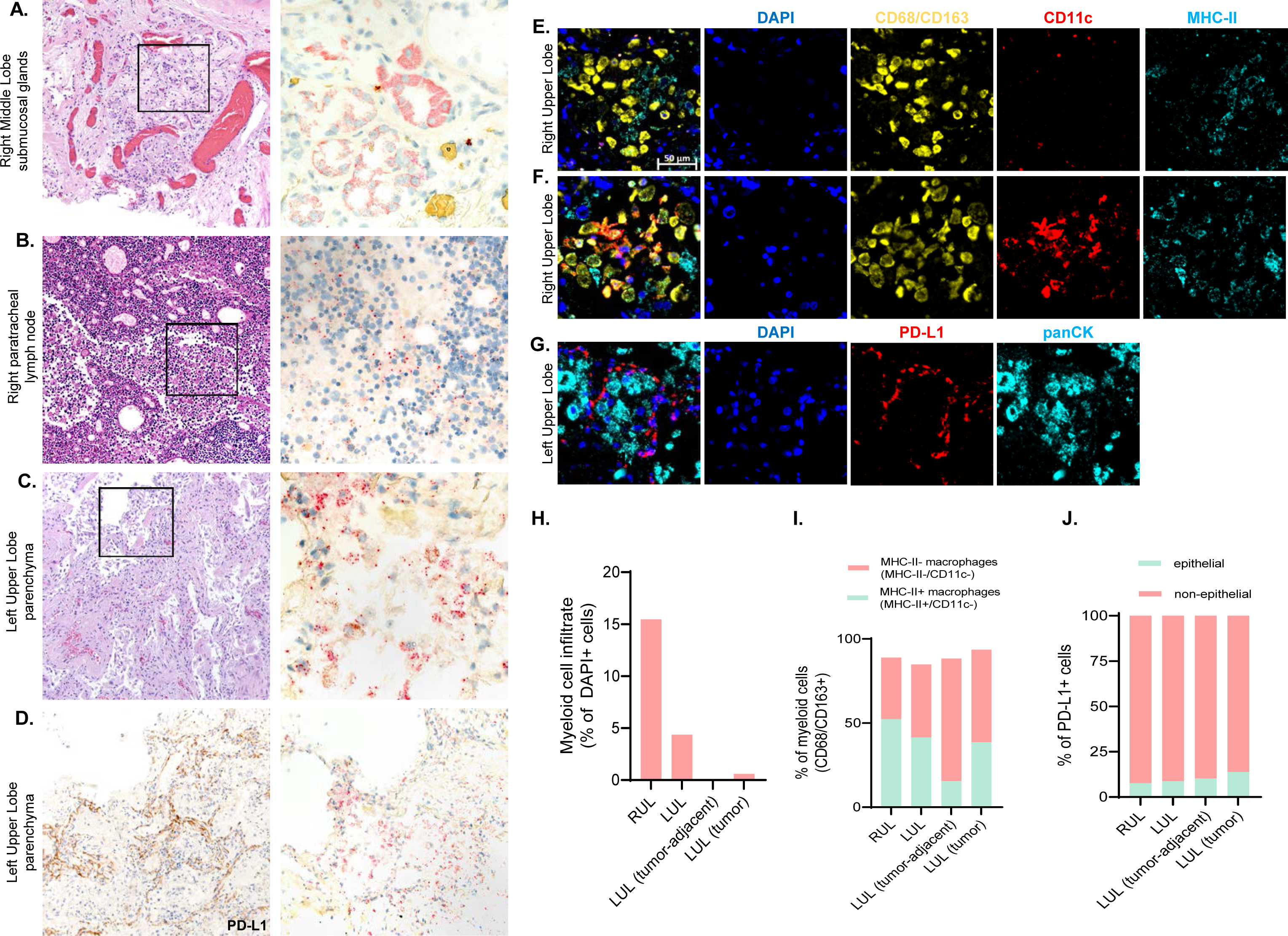
Localization of viral RNA in autopsy tissues and PD-L1 profile. (**A-C**) Representative H&E images (left panels, 200x magnification) and SARS-CoV-2 RNA by RNAscope™ (right panels, 200x magnification, viral RNA stained red) for the following anatomical locations: (**A**) a submucosal gland in the right middle lobe of the lung, (**B**) right paratracheal lymph node, and (**C**) parenchyma from the left upper lobe of the lung. (**D**) PD-L1 staining (left panel, 200x) and RNAscope™ (right panel, 200x) for the left upper lobe of the lung. Multiplex immunofluorescence labeling was performed with the Ultivue UltiMapper™ I/O PD-L1 and UltiMapper™ I/O APC kits. Representative images (20x magnification) from the right upper lobe (**E**, **F**) and left upper lobe (**G**) are shown. Scale bar: 50 μm. In panel (**E**), CD68+/CD163+ myeloid cells and MHC-II+ cells occupy the alveolar space. In panel (**F**), the CD68+/CD163+ myeloid cells in the alveolar space co-express CD11c and/or MHC-II. In panel (**G**), the alveolar space is occupied by panCK+ epithelial cells. PD-L1 is expressed on non-epithelial cells in alveolar walls. (**H**) Quantification of the myeloid cell infiltrate per sample. (**I**) Quantification of MHC-II expression in the myeloid cell infiltrate per sample. (**J**) Quantification of panCK expression in PD-L1 expressing cells per sample. RUL: right upper lobe. LUL: left upper lobe. Tumor-adjacent tissue showed no signs of acute lung injury.

Sections of the primary tumor in the left upper lobe consisted of extensive areas of tumor necrosis with small islands of viable tumor on the periphery of the mass (**Fig. 2C**). The tumor was infiltrated by CD68+ and CD3+ cells, with a predominance of CD68+ cells. The extensive tumor necrosis observed is consistent with the reduction in the size of the primary tumor mass observed on CT scans after one cycle of therapy (**Fig. S2A**). In addition, nuclear material indicative of acute tumor lysis syndrome was observed in small vascular spaces on hematoxylin and eosin stained sections, and the presence of nucleic acid in the vessel lumens was confirmed using DAPI (**Fig. S2B**).

Utilizing RNA *in situ* hybridization, SARS-CoV-2 viral RNA was identified in the submucosal glands of a large airway (**Fig. 3A**), in macrophages in a paratracheal lymph node (**Fig. 3B**), and in the pulmonary interstitium (**Fig. 3C**). In the interstitium, viral RNA was observed in intra-alveolar macrophages, alveolar walls, and sloughed epithelial cells (**Fig. 3C**). There was increased PD-L1 signal in alveolar walls proximal to regions where viral RNA was detected (**Fig. 3D**) compared to regions of the lung where virus was not detected (**Fig. S3A,B**).

Multiplexed staining of the pulmonary interstitium demonstrated that the alveolar space was occupied by CD68+/CD163+ myeloid cells (**Fig. 3E**) and panCytokeratin positive epithelial cells (sloughed pneumocytes, **Fig. 3G**). Consistent with our previous observations, the right lung had more extensive myeloid cell infiltrate (**Fig. 3H)**. Most CD68+/CD163+ myeloid cells co-expressed MHC-II and were predominantly CD11c negative (**Fig. 3F,I**), consistent with pro-inflammatory macrophages. PD-L1 was mainly expressed by non-epithelial cells (**Fig. 3J**) histologically identified as endothelial cells and mononuclear myeloid cells (**Fig. 3G**).

Finally, microscopic evaluation of the right ventricle, left ventricle, and intraventricular septum of the heart was unremarkable with no evidence of acute myocarditis or myocardial degeneration (data not shown).

## Discussion

Cancer patients represent a uniquely vulnerable population during the COVID-19 pandemic^2,3^. Here, we describe the case of a patient with newly diagnosed lung cancer who developed rapidly fatal pneumonitis after his first dose of chemotherapy plus immunotherapy. The initial presenting symptoms (dyspnea, hypoxemia, and cough without fever) and radiographic findings were attributed to ICI-pneumonitis from atezolizumab. However, he failed to improve with standard treatment for ICI-pneumonitis (high dose steroids, infliximab) and was found to have SARS-CoV-2 infection by a nasal swab RT-PCR assay. Post-mortem, blood samples obtained prior to the positive nasal swab result indicated the presence of an IgG response to SARS-CoV-2.

The temporal dynamics of IgG response to SARS-CoV-2, which initial data suggest occur at a median of 13 days after symptom onset with 100% of patients achieving seroconversion by 20 days^4^, suggest that this patient may have been exposed early in his clinical course. Importantly, IgG and IgM titers were above background and quantifiable, but lower than some of the PCR-positive controls (range 5-25 days post-symptom onset). The implications of these low-positive results are not completely clear.

In a recent small case series of cancer patients with SARS-CoV-2 infection, patients who had received ICI therapy had higher risk of death and severe symptoms^5^. ICI therapies targeting the PD-1/PD-L1 axis are standardly used for the treatment of numerous malignancies. These therapies function to activate the immune system by inhibiting T cell exhaustion and tolerance. However, by targeting cancer-induced immunosuppression, ICI therapies can also disrupt immunologic self-tolerance leading to immune-related adverse events (irAEs) in multiple organs, including ICI-pneumonitis^6^. Furthermore, the PD-1/PD-L1 axis, through which ICI therapies exert their anti-tumor response, also regulates anti-viral responses, and upregulation of PD-1 and PD-L1 has been observed during viral infection^7^. Overall, the complex interplay between SARS-CoV-2 infection, resultant viral induced inflammation, and the immune modulating effects of ICI therapies used to treat cancer are not well understood.

This case illustrates several important challenges related to treatment of cancer during the COVID-19 pandemic. First, symptoms of SARS-CoV-2 infection may overlap with symptoms from cancer itself and toxicities of anti-cancer therapies. In this case, the patient was initially diagnosed with ICI-pneumonitis, which is a severe and potentially life-threatening complication that typically presents with progressive dyspnea, hypoxemia, and cough with imaging findings and histopathological changes that overlap with those of other infectious and inflammatory conditions, including COVID-19^6,8^. Both ICI pneumonitis and COVID19 infection can also result in fulminant clinical presentations, including acute respiratory distress syndrome (ARDS). While the clinical and radiographic presentations between COVID-19 infection and ICI pneumonitis are non-specific and overlap, the treatments are different. High-dose steroids with prolonged taper are the mainstay of treatment for ICI-pneumonitis. However, a recent meta-analysis suggested that steroids may cause harm in cases of viral pneumonia and ARDS from influenza^9^. Moreover, ICI-pneumonitis responds to TNFα blocking agents (e.g., infliximab), while the efficacy of this agent in the setting of COVID-19 infection is unknown. Differentiating between these possible causes of respiratory symptoms is challenging yet necessary to initiate the correct treatment.

This case also illustrates knowledge gaps in our understanding of how the immune system responds to SARS-CoV-2 infection in the setting of concurrent cancer immunotherapy. Through examination of autopsy specimens, it appeared that administration of immunotherapy may have caused a profound lung-specific immunologic reaction which may have been responsible for the patient’s death. Serologic tests were positive for an IgG response to the SARS-CoV-2 virus from blood taken prior to the nasal swab viral RT-PCR assay. Though the timing of the initial viral infection cannot be precisely determined, the presence of an IgG response suggests that the viral infection occurred before the patient received any cancer-directed therapy. This timeline for viral infection is supported by the histopathological changes observed in the lungs. In particular, there were elements of both acute pneumonia as well as organizing pneumonia. In this case, we hypothesize that the typical inflammatory response to viral infection was intensified by the administration of a PD-L1 inhibitor, leading to a surge of cytokines / chemokines that overwhelmed the patient’s normal homeostatic processes and led to severe lung injury / pneumonitis and ultimately death from ARDS. This hypothesis is supported by the brisk progression of his respiratory symptoms after ICI-therapy, the rapid radiographic evolution of pneumonitis on his CT scans obtained at close intervals despite standard treatment for ICI-pneumonitis, and the presence of both viral RNA in mucosal surfaces as well as evidence of an IgG response to the virus.

Finally, this case underscores that the optimal approach for viral testing and treatment in relation to the timing of anti-cancer therapies during this global pandemic remains unclear. Should COVID-19 status be assessed in advance of starting any new cancer directed therapy or specifically prior to administration of ICIs or highly immunosuppressive regimens? Should therapy be withheld in patients with active COVID-19, and if so, for how long? Is there a certain titer of anti-SARS-CoV-2 antibody required to prevent viral reactivation in the face of systemic therapy for cancer? With the availability of more widespread testing for acute viral infection and serologic assays to evaluate for host anti-viral responses, such evaluation becomes increasingly feasible. At our institution, cancer patients receiving the most immunosuppressive chemotherapy regimens are now routinely tested for SARS-CoV-2 in advance of starting therapy. Furthermore, it is not known if or how ICI-therapies affect viral immunity. Recently, cases of COVID-19 reactivation in non-cancer patients have been described^10^. It is possible that release of immune tolerance through administration of a PD-1/PD-L1 inhibitor could permit viral reactivation, or alternatively, cause an unrestrained immunologic attack on a smoldering viral infection.

In conclusion, through a concerted multi-disciplinary effort, we provide detailed clinical, radiographic, and histopathological evidence to document the complex interplay between SARS-CoV-2 infection, underlying lung cancer, and activation of host immunity by ICI-therapy. Future prospective clinical trials with robust integrated biomarker studies will provide additionally needed insights regarding the risks associated with immunotherapy treatments for cancer patients in the context of concurrent or prior SARS-CoV-2 infection, leading to better strategies to treat both the viral infection and the cancer.

## Data Availability

Reviewed the CARE Checklist of information to include when writing a case report. For item #12 in the checklist “The patient should share their perspective in one to two paragraphs on the treatment(s) they received” -- our case report is about a deceased patient, so we were unable to obtain the patient’s perspective on the treatment received.

## Acknowledgements

First and foremost, the investigator wish to thank the patient and his wife for their courage and generosity. The investigators are very grateful for assistance from the team in the Translational Pathology Shared Resource facility, especially Karen Thompson and Frances Shook, and to Amy Moore, Portia Thomas, and Darren Tyson for reviewing the manuscript.

---

**Supplementary Figure 1:**
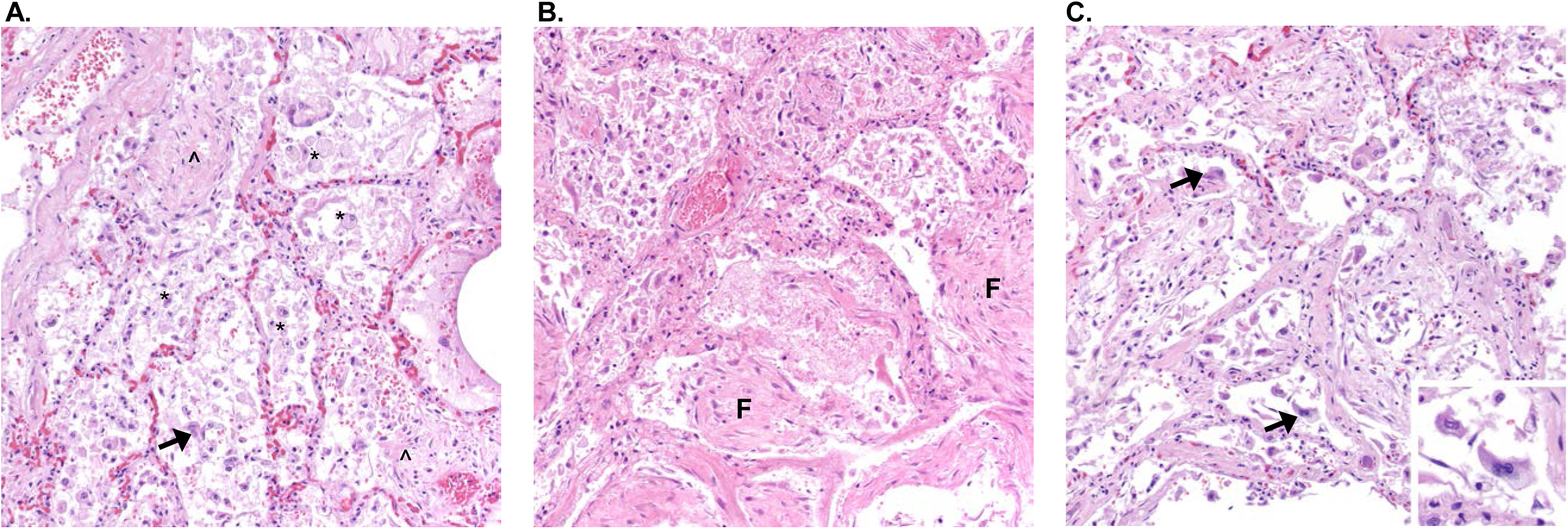
Representative images of lung with diffuse organizing alveolar damage. H&E stain, 200x magnification, Images of right lung showing (**A**) Acute lung injury with foamy macrophages (*), sloughing epithelial cells (arrow), fibromyxoid-organizing exudates (^), fibrin, hemorrhage, and edema. (**B**) Mature fibrosis and fibrotic nodules (F: fibrosis); (**C**) Sloughed reactive cells(arrow). Inset: Digitally magnified reactive cells with amphophilic cytoplasm, enlarged nuclei, multi-nucleation, and nucleoli.

**Supplementary Figure 2:**
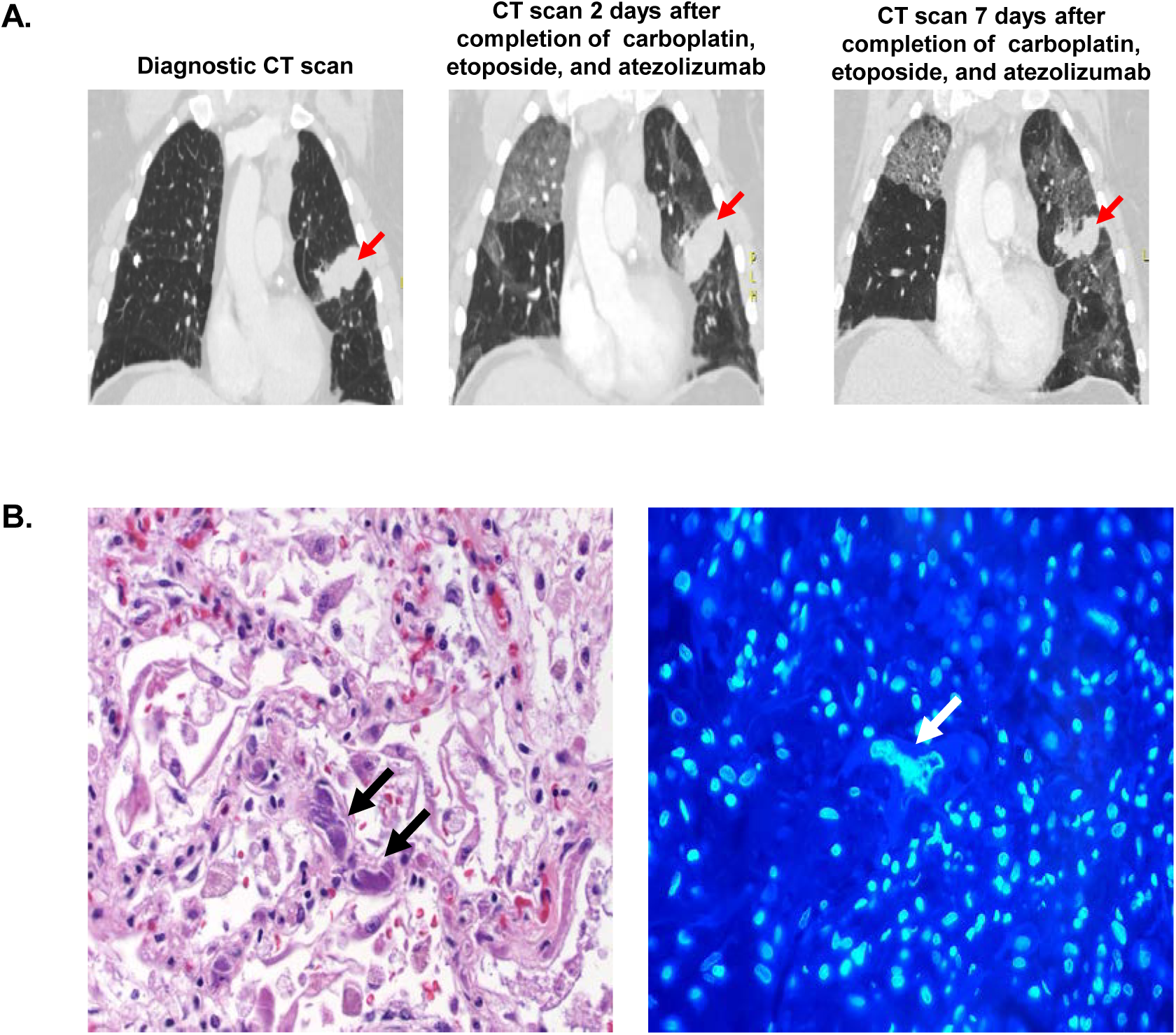
Dramatic anti-tumor response within one week of administration of cycle 1 of chemotherapy plus immunotherapy. (**A**) CT scans showing tumor reduction from the initial diagnostic scan (left panel) to the scans obtained 2 days (middle panel) and 7 days (right panel) after completion of carboplatin, etoposide, and atezolizumab. Red arrow shows tumor mass in the left lung. (**B**) Sections of a vessel in the right upper lobe were stained with H&E (left panel) and DAPI (right panel). Black arrows on the H&E image depict chromatin thrombi. White arrows on the DAPI image depict nucleic acid in the vessel.

**Supplementary Figure 3:**
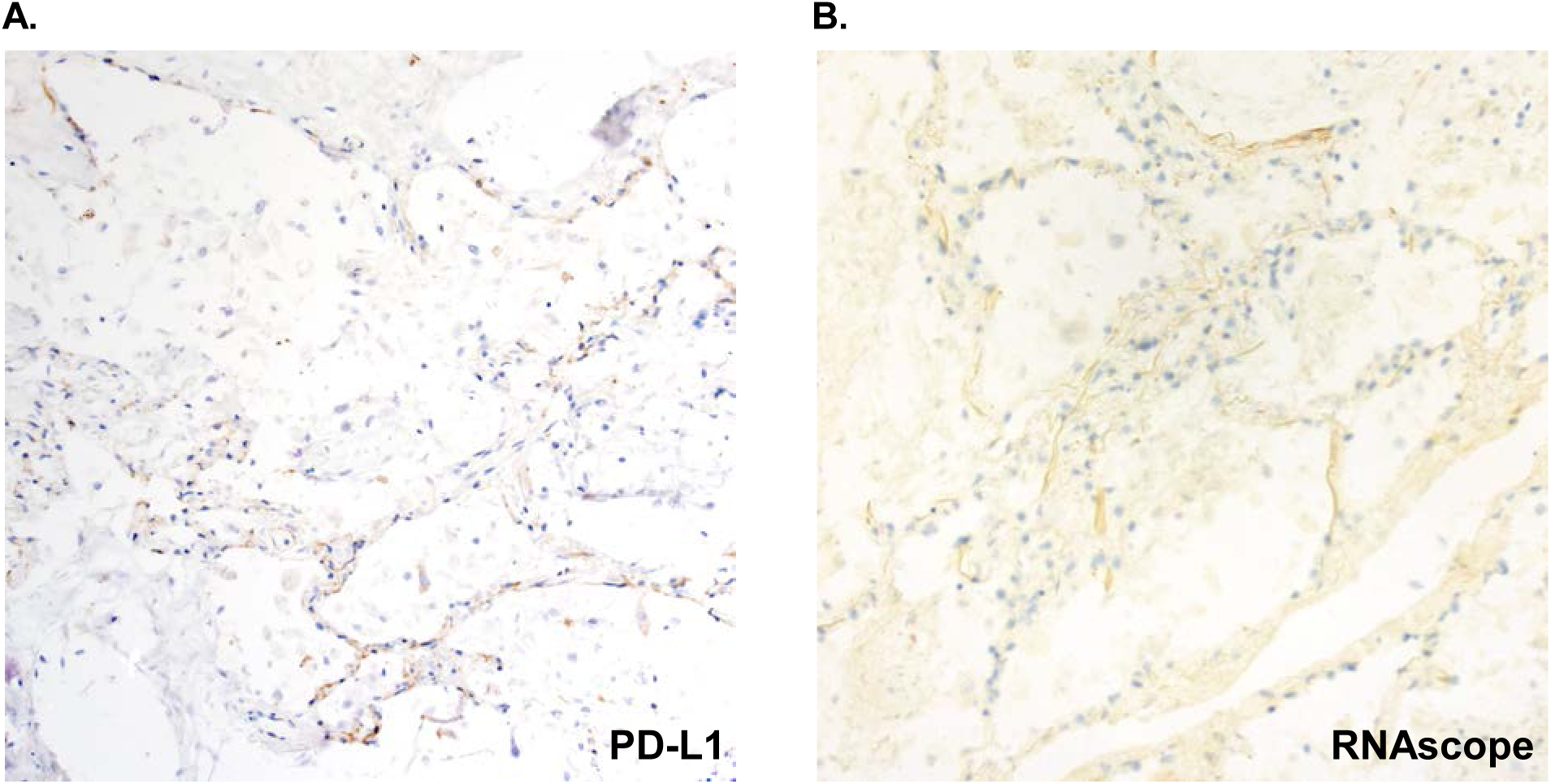
Localization of viral RNA in tissues and PD-L1 profile. PD-L1 staining, 200x magnification (**A**) and RNAscope™ (viral RNA stained red), 200x magnification (**B**) for the right upper lobe of the lung.

## References

1. Bankhead P, Loughrey MB, Fernandez JA, et al. QuPath: Open source software for digital pathology image analysis. Scientific reports 2017;7:16878.

2. Wu Z, McGoogan JM. Characteristics of and Important Lessons From the Coronavirus Disease 2019 (COVID-19) Outbreak in China: Summary of a Report of 72314 Cases From the Chinese Center for Disease Control and Prevention. Jama 2020.

3. Zhou F, Yu T, Du R, et al. Clinical course and risk factors for mortality of adult inpatients with COVID-19 in Wuhan, China: a retrospective cohort study. Lancet 2020;395:1054–62.

4. Long Q-x, Deng H-j, Chen J, et al. Antibody responses to SARS-CoV-2 in COVID-19 patients: the perspective application of serological tests in clinical practice. medRxiv 2020:2020.03.18.20038018.

5. Dai M-YL, Dianbo; Liu, Miao; Zhou, Fu-Xiang; Li, Gui-Ling; Chen, Zhen; Zhang, Zhi-An; You, Hua; Wu, Meng; Zhen, Qi-Chao; Xiong, Yong; Xiong, Hui-Hua; Wang, Chun; Chen, Chang-Chun; Xiong, Fei; Zhang, Yan; Peng, Ya-Qin; Ge, Si-Ping; Zhen, Bo; Yu, Ting-Ting; Wang, Ling; Wang, Hua; Liu, Yu; Chen, Ye-Shan; Mei, Jun-Hua; Gao, Xiao-Jia; Li, Zhu-Yan; Gan, Li-Juan; He, Can; Li, Zhen; Shi, Yu-Ying; Qi, Yu-Wen; Yang, Jing; Tenen, Daniel G.; Chai, Li; Mucci, Lorelei Ann; Santillana, Mauricio; Cai, Hong-Bing. Patients with Cancer Appear More Vulnerable to SARS-CoV-2: A Multi-Center Study During the COVID-19 Outbreak The Lancet 2020;Available at SSRN: http://dx.doi.org/10.2139/ssrn.3558017.

6. Naidoo J, Wang X, Woo KM, et al. Pneumonitis in Patients Treated With Anti-Programmed Death-1/Programmed Death Ligand 1 Therapy. J Clin Oncol 2017;35:709–17.

7. Schonrich G, Raftery MJ. The PD-1/PD-L1 Axis and Virus Infections: A Delicate Balance. Front Cell Infect Microbiol 2019;9:207.

8. Sears CR, Peikert T, Possick JD, et al. Knowledge Gaps and Research Priorities in Immune Checkpoint Inhibitor-related Pneumonitis. An Official American Thoracic Society Research Statement. Am J Respir Crit Care Med 2019;200:e31-e43.

9. Ni YN, Chen G, Sun J, Liang BM, Liang ZA. The effect of corticosteroids on mortality of patients with influenza pneumonia: a systematic review and meta-analysis. Crit Care 2019;23:99.

10. Yuan J, Kou S, Liang Y, Zeng J, Pan Y, Liu L. PCR Assays Turned Positive in 25 Discharged COVID-19 Patients. Clin Infect Dis 2020.

